# Traveler-based Genomic Surveillance: A Scalable Approach to Early Pathogen Detection and Global Biosecurity

**DOI:** 10.64898/2026.04.28.26351949

**Authors:** Stephen M. Bart, Teresa C. Smith, Andrew P. Rothstein, Grace D. Appiah, Samantha M. Loh, Dawn Gratalo, Birgitte B. Simen, Casandra W. Philipson, Robert C. Morfino, Sarah Anne J. Guagliardo, Ian Ruskey, Allison Taylor Walker, Patti Ward, Ezra T. Ernst, Daniel C. Payne, Martin S. Cetron, Cindy R. Friedman

## Abstract

**Background:** In September 2021, the U.S. Centers for Disease Control and Prevention (CDC) implemented the Traveler-based Genomic Surveillance (TGS) program, a surveillance system that leverages genomic sequencing of samples from international air travelers and aviation wastewater for early detection of infectious threats.

**Methods:** During September 2021–August 2024, nasal samples were collected anonymously from volunteer international travelers arriving at eight U.S. airports. During February 2023– August 2024, aviation wastewater samples were collected from arriving flights. Nasal samples were pooled and sent to a laboratory for RT-PCR testing. Genomic sequencing was conducted for SARS-CoV-2 and respiratory, gastrointestinal (wastewater), and other pathogens of public health importance.

**Findings:** Nasal samples from 694,798 travelers were grouped into 67,308 pools and tested; 13,990 (20.8%) were positive for SARS-CoV-2. Over 80% (400/495) of airplane and 96·6% (422/437) triturator (a wastewater collection point from multiple airplanes) samples were positive for SARS-CoV-2. Sequence results were made publicly available a median of 11 days (IQR 10– 13 days) after sample collection. Predominant SARS-CoV-2 variants changed over time. Positive tests for influenza virus and respiratory syncytial virus were high in December/January, and gastrointestinal viruses were detected in wastewater during all months. Monitoring was scaled in response to reported outbreaks of COVID-19 and *Mycoplasma pneumoniae* in China and clade 1 monkeypox virus in central Africa.

**Interpretation:** Traveler nasal and aviation wastewater sampling can provide critical early detection of infectious pathogens before widespread U.S. community transmission. The TGS program provides a model for integrated traveler-based genomic surveillance.

**Funding:** CDC

**Research in Context:** *Evidence before this project:* We searched PubMed for relevant studies published during December 1, 2020–August 31, 2024, using the terms “traveler surveillance”, “wastewater monitoring”, “SARS-CoV-2 genomics”, and “airport-based surveillance”, without language restrictions. Previous reports have shown the feasibility of using travelers as sentinel populations for disease surveillance. Modeling studies have proposed integrating genomic data into international travel surveillance systems to enhance early pathogen detection, and evidence from Australia, Canada, and the UK suggests such programs could be scalable and effective. Early pandemic-era wastewater surveillance, particularly aviation wastewater, demonstrated that air travel hubs can be used to monitor pathogen importation. Prior efforts largely focused on SARS-CoV-2, with limited integration of multi-pathogen surveillance or side-by-side comparisons of nasal and wastewater surveillance modalities. A limited number of public health reviews have examined the broader implications of airport-based surveillance, including novel methods like airplane wastewater testing. However, empirical data on sustained, large-scale implementation of these models especially outside of regulatory or mandatory testing frameworks have been sparse.

*Added value of this project:* This is the first real-world implementation and scale-up of an anonymized, multi-pathogen traveler-based surveillance system across multiple U.S. international airports. We developed a scalable framework that integrated nasal swab testing, airplane and airport wastewater sampling, with genomic sequencing into a unified pathogen surveillance platform. Unlike prior efforts which primarily focused on SARS-CoV-2, this program captured respiratory and gastrointestinal viruses simultaneously and tracked genomic variation in near-real time. The program transitioned from a pilot to a multi-modality national surveillance system in under four years, engaging nearly 700,000 international travelers, and nearly 1000 aviation wastewater samples. Our findings demonstrate the feasibility of rapidly adapting this infrastructure for emerging threats and underscores the importance of sentinel surveillance in addressing global sequencing blind spots.

*Implications of all the available evidence:* The successful scale-up and real-time application of the TGS program illustrates that traveler-based surveillance can serve as a critical global early warning tool. Data generated from this program have filled gaps in global pathogen tracking, informed public health responses to outbreaks, and demonstrated that surveillance of international travelers can be achieved without mandatory testing. The scalability, speed, and adaptability of the program offer a viable model for global replication, especially as routine surveillance capacities decline. Our findings suggest that integration of multi-modal, voluntary traveler surveillance including sequencing and wastewater-based epidemiology should be considered a core component of pandemic preparedness and response frameworks worldwide.

## Background

Travelers can rapidly spread pathogens across borders, making them an important population for tracking emerging infectious diseases. Understanding which pathogens and genomic variants are entering a country—before they spread locally—can guide rapid public health actions and help avoid wider disruptions to travel and trade.

For decades, international travel clinic-based sentinel surveillance has helped identify new and emerging global infectious disease trends.^1^ During the COVID-19 pandemic, however, case detection through clinic-based sentinel surveillance was hindered by the mild or asymptomatic nature of many infections and the widespread use of at-home COVID-19 tests.^2^ Global SARS-CoV-2 surveillance has relied on laboratory reporting to public health and public genomic sequence submissions, but since January 2022, both testing and sequencing have declined 95% worldwide, with marked geographic variation in reporting and timeliness.^3,4^

The U.S. Centers for Disease Control and Prevention (CDC) launched the Traveler-based Genomic Surveillance (TGS) program in 2021 to detect SARS-CoV-2 variants entering the United States before widespread transmission. During an initial eight-week pilot, we collected nasal samples from volunteer travelers arriving from India at three U.S. airports and tested only for SARS-CoV-2.^5^ The program later expanded to additional airports, pathogen targets, and travelers from many countries. Following a 2022 pilot, TGS also began sampling airplane wastewater for respiratory, gastrointestinal, and other pathogens, providing a modality that does not require direct traveler participation.^6^ TGS is designed to be nimble and scalable, with goals that include early detection of genetic changes in pathogens of public health importance, monitoring trends, and filling gaps in global pathogen surveillance.

Traveler nasal swab surveillance and aviation wastewater surveillance involve different levels of traveler engagement and yield data with varying granularity. Nasal swab sampling relies upon volunteer traveler participation, yielding a heterogeneous sample of travelers from selected flights, and allows collection of health information and travel details for each participant, although participation is anonymous. Aviation wastewater surveillance, conversely, does not require individual travelers’ opting in, and collection yields a composite sample from many travelers on a flight without traveler-level metadata, although it requires cooperation from airlines and airports and reflects only those travelers who use the airplane lavatory.^7^ Aviation wastewater samples can also be collected from downstream sites, such as the airport triturator—where waste from multiple flights is discharged and homogenized, before later being combined with airport terminal wastewater.^8^

Data from the TGS program have been used for early variant detection,^5,9^ evaluation of public health interventions,^10^ and addressing gaps in global surveillance.^11^ In this report, we describe TGS data collected from 2021 – 2024 and highlight its utility for international pathogen surveillance.

## Methods

### Program overview

The Traveler-based Genomic Surveillance program is a public-private partnership between CDC, Ginkgo Biosecurity, and XpresCheck, established to monitor emerging and circulating pathogens among international air travelers arriving in the United States.^12^

### Nasal swab sampling and testing

International travelers aged ≥18 years were invited to participate voluntarily at selected U.S. airports, regardless of symptoms. After providing consent, participants self-collected anterior nasal swab samples and completed a brief questionnaire capturing demographics, travel itinerary, and reasons for participation (e.g. supporting public health or receiving a free at-home self-test kit). Testing began in September 2021 and expanded over time to eight U.S international airports. Until September 2022, participants were also given a mail-in SARS-CoV-2 saliva test kit for use 3–5 days later (**Supplementary Methods**).

Samples were shipped daily to a central laboratory and tested using a pooled reverse transcription-polymerase chain reaction (RT-PCR) testing strategy. During September 2021– January 2023, a single swab was collected from each participant and swabs were grouped into pools (5-25 swabs) by flight-origin country (**Figure 1**). Beginning in late December 2022, two swabs were collected from each participant; one was used for pooled testing and the other held for reflex testing if the pool tested positive for SARS-CoV-2, influenza A or B virus, or respiratory syncytial virus (RSV) (influenza A and B viruses and RSV testing added in 2023). Pooled testing served as a triage step: when a pool was positive, all individual swabs represented in that pool were tested (ThermoFisher TrueMark SARS-CoV-2, Flu A, Flu B, RSV Select Panel). If the pool was negative, all individuals were presumed negative.

During December 2023–March 2024, samples from two airports were also tested for *Mycoplasma pneumoniae* (ThermoFisher TrueMark Respiratory Panel OpenArray) in response to reports of an outbreak in China of *M. pneumoniae* associated with macrolide resistance (Supplementary Methods).^13^

### Wastewater sampling and testing

Airplane lavatory and airport triturator wastewater were collected at selected airports (Supplementary Methods). Airplane wastewater was obtained from the lavatory service port of individual arriving flights while triturator wastewater consisted of 24-hour composite samples collected from multiple flights. Samples were shipped to a central laboratory and tested using RT-PCR (February-December 2023) and RT-digital PCR (RT-dPCR) from December 2023– August 2024, using multiplex panels for SARS-CoV-2 and select respiratory and enteric pathogens (Supplementary Methods).

### Genomic sequencing

All nasal samples (pooled or individual) or wastewater samples positive for SARS-CoV-2 underwent whole genomic sequencing. Influenza virus- and RSV-positive individual nasal samples were also sequenced. Consensus genomes passing quality control criteria were assigned to lineages or subtypes (Supplementary Methods). Wastewater samples, underwent lineage deconvolution to identify multiple SARS-CoV-2 lineages in mixed samples (supplementary methods). Influenza virus-positive samples were monitored for the emerging influenza A(H5N1) and the potentially extinct influenza B/Yamagata lineages.^14,15^ Sequences with notable mutational patterns or with emerging lineages underwent additional characterization at CDC.

### Data analysis

We analyzed samples collected September 29, 2021–August 25, 2024. Traveler origin countries were assigned to World Health Organization (WHO) regions based on either the origin of the flight leg arriving in the United States or the origin country of the traveler’s flight itinerary (Supplementary Table 1). Samples with inconclusive results and those not tested due to shipping delays were excluded from results calculations.

Percent positivity was calculated as the number of positive tests divided by the number of valid (positive and negative) results. For SARS-CoV-2, similar sublineages were aggregated for temporal trend analysis. Individual nasal sample RT-PCR results and lineages were assessed by WHO region to identify surges and differences in circulating variants over time.

Turnaround time was defined as the number of days between sample collection and either the RT-PCR result or GISAID sequence submission.

### Ethics

This activity was reviewed by CDC, deemed research not involving human subjects, and was conducted consistent with applicable federal law and CDC policy (see e.g., 45 C.F.R. part 46; 21 C.F.R. part 56; 42 U.S.C. §241(d), 5 U.S.C. §552a, 44 U.S.C. §3501 et seq).

## Results

### Nasal swab sampling participation

During September 29, 2021–August 25, 2024, 694,798 travelers participated in the TGS program (**Figure 1**). A plurality of participants originated in the European region (34·5%, 240,029 participants) (**Supplementary Table 1**). There were 151 origin countries with ≥50 participants; travelers from India (n = 66,649), the United Kingdom (n = 57,033), Japan (n = 37,303), Mexico (n = 34,441), and the Dominican Republic (n = 34,391) were the most numerous (**Supplementary Figure 1**). Most participants were aged 18–49 years (419,938/630,621 respondents, 66·6%) and U.S. residents (352,361/603,651 respondents, 58.4%). Participants most frequently reported female sex, non-Hispanic ethnicity, and White race (**Supplementary Table 1**). For the primary reason for their travel, most participants reported tourism/vacation (384,072/568,380 respondents, 67·5%), visiting friends/relatives (99,774, 17·6%), or a business/occupational reason (62,900, 11·1%).

When asked why they participated, for which they could pick multiple reasons, 70·6% of participants (398,951 of 565,122 respondents) responded that they wanted to support public health work, help the CDC monitor diseases entering the United States, or both; 47·5% (268,925) wanted the free at-home self-test kit.

### Nasal swab and saliva sampling test results and sequencing for SARS-CoV-2

Among 67,308 pooled samples tested during September 2021–August 2024, 13,990 (20·8%) were positive for SARS-CoV-2, with weekly percentages ranging 5.6–42·9% (**Figure 2A**).

Pooled test results were available aproximately 36 hours after collection, including an average of 28 hours for shipping time. Of 6,299 resulted at-home saliva samples, 546 (8·7%) were positive. Trends in the proportion of positive at-home saliva samples mirrored those for positive pooled tests before at-home sample collection was discontinued (**Supplementary Figure 2A and 2B**).

During December 2022–January 2023, when each traveler contributed two samples, individual test results were available approximately 62 hours after collection. This longer turnaround time reflected the need to first complete pooled testing before testing the individual swabs from positive pools. Among 570,889 travelers, 15,655 individual samples (2·7%) were positive for SARS-CoV-2, with weekly percentages ranging 1.1–5·1%. Trends in individual tests closely mirrored those observed in pooled testing (**Figure 2A, inset**).

SARS-CoV-2 sequencing results were published in GISAID a median of 11 days (IQR 10–13 days) after sample collection. Samples lineages differed by year (Figure 2B) and were predominantly the Delta in late 2021; Omicron BA.1, BA.2, BA.4/BA.5, and BQ.1 in late 2021 through 2022; Omicron XBB, XBB.1.5, XBB.1.9, XBB.1.16, and EG.5 in 2023; and Omicron BA.2.86/JN.1, KP.2, and KP.3 in late 2023 through 2024 (**Figure 2B**).

Test results and lineages from individual samples were also assessed (**Supplementary Figure 3**). Noted patterns included increases in percentages of positive test results associated with XBB lineages in travelers from the Eastern Mediterranean and Southeast Asian regions in early 2023 and a wave of EG.5 infections in arriving European travelers in September–October 2023.

### Nasal swab sampling test results and sequencing for influenza viruses and RSV

During October 2023–August 2024, 203,449 samples from travelers were tested for influenza A virus, influenza B virus, and RSV, of which 513 (0.3%) were positive for influenza A virus; 53 (0·03%) for influenza B virus; and 1,097 (0·6%) for RSV (**Figure 3A**). The proportion of positive influenza A virus and RSV test results peaked in late December 2023.

Overall, slightly more influenza A viruses detected were of the H3N2 subtype (229/421 subtyped influenza A viruses, 54·4%) than H1N1 (192/421, 45·6%); H5N1 was not detected (**Figure 3B**). The H3N2 subtype was the predominant serotype detected during April–August 2024 (129/210 subtyped influenza A viruses, 61·4%) and was most often detected in travelers from Central and South America (data not shown). All sequenced influenza B viruses (n = 35) were of the Victoria subtype; influenza B/Yamagata was not detected (**Figure 3C**). The majority of RSV-positive samples sequenced identified RSV B (98/155, 63·2%) (**Figure 3D**).

### Airplane wastewater surveillance for SARS-CoV-2

During March 2023–August 2024, 499 airplane wastewater samples from flights arriving from one Eastern Mediterranean country were tested for SARS-CoV-2; four had inconclusive results (**Figure 4A**). Most samples were positive (400/495, 80·8%) (**Figure 4B**). Lineages were identified in 259 (64·8%) positive samples, including 104 (26·0%) positive samples that had multiple lineages identified. Lineage proportions from airplane wastewater (**Figure 4C**) were comparable with those from contemporaneous Eastern Mediterranean region nasal swab samples (**Supplementary Figure 3C**), though some variants were identified in nasal sampling before wastewater.

### Triturator wastewater surveillance for SARS-CoV-2

During April 2023–August 2024, 443 triturator wastewater samples were collected; six had inconclusive results. Almost all triturator samples were positive (422/437 samples, 96·6%) (**Figure 4B**). Lineages were identified in 376 (89·1%) positive triturator samples including 285 (67·5%) where multiple lineages were identified, reflecting a greater proportion of samples with multiple lineages than airplane wastewater. Lineage proportions from triturator detections (**Figure 4D**) were comparable with those observed from nasal swab sampling during the same time (**Figure 2B**).

### Airplane and triturator wastewater surveillance for other pathogens

During December 2023–August 2024, airplane (n = 294) and triturator (n = 248) wastewater samples were tested for an expanded panel of respiratory viruses and gastrointestinal viruses. Respiratory (influenza A and B viruses, RSV) and gastrointestinal (norovirus, adenovirus F) viruses were commonly detected in both airplane and triturator wastewater (**Figure 5**). Respiratory virus detections were highduring December 2023–January 2024 but continued at a lower level after the U.S. respiratory virus season ended. Gastrointestinal viruses did not exhibit obvious seasonality but were frequently detected in both airplane and triturator wastewater.

### COVID-19 surge in China during December 2022–January 2023 and other public health events

Beginning December 7, 2022, China relaxed its “zero COVID” policy, leading to a surge of COVID-19 cases.^16^ During this surge, there were significant gaps in public data regarding cases, hospitalizations, and deaths.^17^ A major concern was the potential for emergence of a new variant that could spread internationally, but few sequences were initially submitted into international databases. To fill these gaps, the TGS program extended operating hours at participating airports (especially in San Francisco) and began operations at airports in Seattle and Los Angeles to rapidly increase recruitment of volunteer travelers originating in China and other countries in Eastern Asia (**Figure 6A and Figure 6B**).

The rate of positive tests among TGS program participants originating in mainland China (excluding the Hong Kong and Macau Special Administrative Regions) preceded the increase (and subsequent decrease) in case counts reported to WHO by about one week (**Figure 6C**). During December 7, 2022–January 5, 2023, SARS-CoV-2 was detected in 21 of 115 samples (18·2%) from TGS program participants originating in China. In response to the outbreak, during January 5–March 10, 2023, travelers from China were required to show a recent negative test result or evidence of recent recovery from COVID-19 before boarding a flight to the United States.^18^ During January 5–March 10, SARS-CoV-2 was detected in only four TGS program participants originating in China (4/425, 0·8%), and three of these detections occurred for travelers arriving on January 5 who likely boarded before the policy came into effect. Most sequences from TGS program participants arriving from China during December 2022–January 2023 were BA.5.2 (56·3%, 9/16) or BF.7 (37·5%, 6/16).

Other examples of the TGS program flexing to timely public health events included outbreaks of *M. pneumoniae* and monkeypox virus. In response to the 2023 *M. pneumoniae* outbreak in China, nasal swab samples were tested (n = 2,398). *M. pneumoniae* was detected in samples from two travelers arriving from South America; no positive samples were detected in travelers arriving from China or East Asia. Neither isolate was macrolide-resistant. During December 2023–August 2024, in response to a 2024 clade I monkeypox virus outbreak in Central Africa,^19^ airplane and triturator wastewater was tested using the non-variola orthopoxvirus (NVO) test; no samples tested positive (n = 542) (**Figure 5G and Figure 5N**).

## Discussion

The TGS program is the first real-world large-scale implementation of an anonymized, multi-pathogen, multi-modal, genomic surveillance system across multiple U.S. international airports. We developed a scalable framework that integrates nasal swab testing, aviation wastewater sampling, and genomic sequencing into a unified pathogen surveillance platform. This system was able to detect and track SARS-CoV-2 variants at near-real-time. Unlike earlier initiatives that focused solely on SARS-CoV-2, TGS adopted a multimodal strategy to monitor a broader range of respiratory and gastrointestinal viruses simultaneously. In less than three years, the TGS program engaged nearly 700,000 international travelers who voluntarily and anonymously contributed nasal samples. Unlike mandatory airport testing programs in other countries, TGS demonstrated that a fully voluntary, large-scale pathogen surveillance system can be both feasible and publicly acceptable, and capable of being adopted elsewhere.^20^

Traditional clinical surveillance often misses early signals because it captures only a fraction of infections, typically from symptomatic patients who seek care, and are tested. Traveler-based surveillance, including aviation wastewater sampling, fills gaps by capturing populations and pathogens that traditional systems may not reach. This allows for earlier detection of emerging variants and transmission trends, especially from countries with limited genomic surveillance capacity. Despite the end of the COVID-19 emergency in May 2023, SARS-CoV-2 continues to evolve, with new variants emerging globally while testing and sequencing have sharply declined worldwide. TGS mitigates these blind spots by generating timely, high-quality genomic data, including for pathogens that may not be sequenced through other surveillance systems. Pathogens in samples collected through the TGS program have been forwarded to CDC laboratories for culture, enabling detailed characterization of viral susceptibility to immunity generated from contemporary vaccines.^21^

Aviation wastewater surveillance complements community wastewater programs. While community programs can track local pathogen trends, they may lag in identifying novel variants emerging abroad. Aviation wastewater samples, collected either directly from airplanes or from airport triturators, provide a global early warning signal, especially when paired with nasal sampling of travelers for more detailed data. International consortia are now working to standardize aviation wastewater methods and establish global sentinel networks, underscoring its importance for pandemic preparedness.^22^

TGS has repeatedly demonstrated its ability to pivot in response to emerging threats and global health events. During the COVID-19 surge in China in December 2022–January 2023, when little data were available from within the country, the TGS program expanded operations to collect samples from travelers arriving from the region and delivered variant data in near-real-time. The detection of BA.5.2 and BF.7 Omicron variants in TGS samples aligned with eventual reported community-detected sequences from China in public databases, filling a critical global health intelligence gap.^23–25^ Through testing conducted in response to the reported 2023 macrolide-resistant *M. pneumoniae* outbreak in China, the TGS program identified two positive samples from travelers arriving from South America, both of which were susceptible to macrolides. This rapid adaptation illustrated the program’s responsiveness to the global re-emergence of a pathogen with antimicrobial resistance potential and was an early source of specimens for monitoring the re-emergence of *M. pneumoniae* in the United States.^26^ Similarly, screening of wastewater for clade I monkeypox virus and nasal swabs for influenza A (H5N1) and influenza B/Yamagata highlight how the TGS platform can quickly incorporate testing for novel or re-emerging pathogens of concern as global conditions change.

Recent findings underscore the continued value of the TGS platform. The program successfully identified variants such as BA.2.86 in international travelers before they were widely detected in U.S. communities^9^ and provided sustained genomic data during a period when global sequencing efforts declined by more than 95% (https://gisaid.org/hcov-19-variants-dashboard/). Airplane and triturator wastewater routinely detect both seasonal and non-seasonal respiratory and gastrointestinal viruses, reinforcing the utility of this complementary surveillance modality. The platform proved capable of detecting both symptomatic and asymptomatic infections through non-invasive or minimal engagement approaches.

The rapid turnaround time for the program—1–3 days for PCR results and 11 days for sequencing reports—enables timely data delivery during outbreaks when case information or sequencing may be otherwise limited. It also can provide actionable data for public health decision-making, such as assessing the impact of predeparture testing on postarrival infections,^10^ informing guidance issued to travelers, and near-real-time monitoring of emerging mutations that may impact vaccines or therapeutics.

Public engagement was a key element of the program’s success. From inception until August 2024, nearly 700,000 travelers participated voluntarily, with many citing a desire to support public health efforts or interest in receiving a complimentary at-home test as motivation. This level of participation highlights the feasibility of implementing large-scale, ethically informed surveillance in an airport setting without significant disruption to travel. The success of TGS offers a model for designing pathogen surveillance systems that can scale responsibly during public health emergencies.

As with any surveillance system, the TGS program has limitations. The nasal swabbing program is voluntary and subject to participation bias though it is unlikely to be biased in any particular direction. While test results have been shown to correlate with community incidence patterns in traveler origin countries,^11^ the TGS program is not designed to produce nationally representative prevalence estimates. Anonymous participation means that TGS cannot be used to identify individual infected travelers and conduct contact tracing. Sampling is not systematic and may exclude travelers arriving at domestic terminals who entered the United States via preclearance facilities (https://www.cbp.gov/travel/preclearance) or at nonparticipating airports. Certain regions such as parts of Africa are underrepresented relative to other regions because of limited direct flight volume. Airplane wastewater surveillance, while valuable for detecting circulating pathogens, cannot be linked to individual travelers and is therefore not useful for tracing of individual infected persons. Detection depends on shedding into wastewater, and not all travelers use the lavatory during flight, particularly on shorter routes.^7^ Similarly, lower nasal swabs may not be prefered sample type for certain pathogens. In this analysis, samples with any RT-dPCR amplification were identified as positive, a framework which might slightly overestimate positivity for low-copy targets. Additionally, aviation wastewater surveillance requires formal agreements and coordination with airlines, airport authorities, and ground handling crews. During the program period, only a limited number of airlines partners participated, constraining geographic coverage. Building sustainable aviation wastewater surveillance will require a whole-of-society approach that includes partnerships across the aviation sector and public health system. Despite these limitations, the program’s multimodal structure enables triangulation of data sources to fill critical surveillance gaps and inform timely public health decision-making.

The TGS program represents a nimble, scalable, real-world traveler- and aviation-based genomic surveillance system that has moved from theory to practice. Its success highlights the value of continued investment in genomic sequencing, wastewater surveillance, and voluntary public participation. As global health threats evolve, the TGS program demonstrates how layered, adaptive systems can close surveillance gaps and strengthen global early-warning capabilities.

## Data Availability

Genomic sequences are available in public databases including GISAID, NCBI GenBank, and NCBI SRA. Individual-level data may be made available with an approved proposal and signed data-sharing agreement; please for more information.

https://www.ncbi.nlm.nih.gov/bioproject/989177

https://gisaid.org/

## Acknowledgments

Program participants; participating airlines and airports; the CDC Port Health Protection Branch, including Jordan Burton, Douglas Weigelt, Sarah Meehan, Helen MacGregor, Meron Tsige, Steve Burchell, Joshua Tchan, Jonathan Georges, Erica Sison, Olubunmi Akingkube, Ellis Cameron-Perry, Kara Adams, Matt Palo, Linda Pimental, Erin Rothney; Lisa Rotz, David Fitter, Lauren Elsberry, Marielle Glynn, Ashley Brown, Kristina Theis, Deena Butryn. We thank all Ginkgo Operations, Sequencing, and Epidemiology team members who contributed to this work, with special acknowledgment to past and present Program Managers: Claire Altieri, Ben H. Rome, Amy Schierhorn, Tim Lyden, Lisa Li, Tom Aichele, and Robert C. Morfino.

## Disclosures

AR, DG, BS, CP, and RM were employed by Ginkgo Bioworks at the time of the study and received Ginkgo Bioworks stocks. EE and PW received XWell stocks. SB, TS, GA, SL, SG, IR, AW, DP, MC, CRF declare no competing interests.

## Funding

The Traveler-based Genomic Surveillance program was funded by CDC contract awards 75D30121C12036 and 75D30122C14933.

## Data Sharing

Genomic sequences are available in public databases including <u>GISAID</u>, NCBI GenBank, and <u>NCBI</u> SRA. Individual-level data may be made available with an approved proposal and signed data-sharing agreement; please for more information.

## Disclaimer

The findings and conclusions in this report are those of the authors and do not necessarily represent the official position of the Centers for Disease Control and Prevention.

## References

1. Wilder-Smith A, Boggild AK. Sentinel Surveillance in Travel Medicine: 20 Years of GeoSentinel Publications (1999-2018). J Travel Med. 2018;25(1). doi:10.1093/jtm/tay139

2. Brown AB, Miller C, Hamer DH, et al. Travel-Related Diagnoses Among U.S. Nonmigrant Travelers or Migrants Presenting to U.S. GeoSentinel Sites - GeoSentinel Network, 2012-2021. Morb Mortal Wkly Rep Surveill Summ Wash DC 2002. 2023;72(7):1–22. doi:10.15585/mmwr.ss7207a1

3. Ohlsen EC, Hawksworth AW, Wong K, et al. Determining Gaps in Publicly Shared SARS-CoV-2 Genomic Surveillance Data by Analysis of Global Submissions. Emerg Infect Dis. 2022;28(13):S85–S92. doi:10.3201/eid2813.220780

4. Brito AF, Semenova E, Dudas G, et al. Global disparities in SARS-CoV-2 genomic surveillance. Nat Commun. 2022;13(1):7003. doi:10.1038/s41467-022-33713-y

5. Wegrzyn RD, Appiah GD, Morfino R, et al. Early Detection of Severe Acute Respiratory Syndrome Coronavirus 2 Variants Using Traveler-based Genomic Surveillance at 4 US Airports, September 2021-January 2022. Clin Infect Dis Off Publ Infect Dis Soc Am. 2023;76(3):e540–e543. doi:10.1093/cid/ciac461

6. Morfino RC, Bart SM, Franklin A, et al. Notes from the Field: Aircraft Wastewater Surveillance for Early Detection of SARS-CoV-2 Variants - John F. Kennedy International Airport, New York City, August-September 2022. MMWR Morb Mortal Wkly Rep. 2023;72(8):210–211. doi:10.15585/mmwr.mm7208a3

7. Jones DL, Rhymes JM, Wade MJ, et al. Suitability of aircraft wastewater for pathogen detection and public health surveillance. Sci Total Environ. 2023;856(Pt 2):159162. doi:10.1016/j.scitotenv.2022.159162

8. Bivins A, Morfino R, Franklin A, Simpson S, Ahmed W. The lavatory lens: Tracking the global movement of pathogens via aircraft wastewater. Crit Rev Environ Sci Technol. 2024;54(4):321–341. doi:10.1080/10643389.2023.2239129

9. Bart SM, Rothstein AP, Philipson CW, et al. Notes from the Field: Early Identification of the SARS-CoV-2 Omicron BA.2.86 Variant by the Traveler-Based Genomic Surveillance Program - Dulles International Airport, August 2023. MMWR Morb Mortal Wkly Rep. 2023;72(43):1168–1169. doi:10.15585/mmwr.mm7243a3

10. Bart SM, Smith TC, Guagliardo SAJ, et al. Effect of Predeparture Testing on Postarrival SARS-CoV-2-Positive Test Results Among International Travelers - CDC Traveler-Based Genomic Surveillance Program, Four U.S. Airports, March-September 2022. MMWR Morb Mortal Wkly Rep. 2023;72(8):206–209. doi:10.15585/mmwr.mm7208a2

11. Smith TC, Bart SM, Loh SM, et al. SARS-CoV-2 Sample Positivity in Travellers Can Predict Community Prevalence Rates: Data from the Traveller-Based Genomic Surveillance Programme. Prepr Lancet. Published online February 9, 2024. doi:10.2139/ssrn.4720735

12. Friedman CR, Morfino RC, Ernst ET. Leveraging a Strategic Public-Private Partnership to Launch an Airport-Based Pathogen Monitoring Program to Detect Emerging Health Threats. Emerg Infect Dis. 2025;31(13):35–38. doi:10.3201/eid3113.241407

13. Li Y, Wu M, Liang Y, et al. Mycoplasma pneumoniae infection outbreak in Guangzhou, China after COVID-19 pandemic. Virol J. 2024;21(1):183. doi:10.1186/s12985-024-02458-z

14. Garg S, Reinhart K, Couture A, et al. Highly Pathogenic Avian Influenza A(H5N1) Virus Infections in Humans. N Engl J Med. 2025;392(9):843–854. doi:10.1056/NEJMoa2414610

15. Caini S, Meijer A, Nunes MC, et al. Probable extinction of influenza B/Yamagata and its public health implications: a systematic literature review and assessment of global surveillance databases. Lancet Microbe. 2024;5(8):100851. doi:10.1016/S2666-5247(24)00066-1

16. Bradsher K, Che C, Chien AC. China Eases ‘Zero Covid’ Restrictions in Victory for Protesters. The New York Times. https://www.nytimes.com/2022/12/07/world/asia/china-zero-covid-protests.html. mDecember 7, 2022. Accessed August 18, 2025.

17. Dyer O. Covid-19: China stops counting cases as models predict a million or more deaths. BMJ. 2023;380:p2. doi:10.1136/bmj.p2

18. Requirements for Negative Pre-Departure COVID-19 Test Results or Documentation of Recovery From COVID-19 for Aircraft Passengers Traveling to the United States From the People’s Republic of China. Federal Register. January 5, 2023. Accessed August 18, 2025. https://www.federalregister.gov/documents/2023/01/05/2023-00080/requirements-for-negative-pre-departure-covid-19-test-results-or-documentation-of-recovery-from

19. WHO Director-General declares mpox outbreak a public health emergency of international concern. August 14, 2024. Accessed October 1, 2024. https://www.who.int/news/item/14-08-2024-who-director-general-declares-mpox-outbreak-a-public-health-emergency-of-international-concern

20. Gashegu M, Muvunyi R, Musabyimana JP, et al. Early detection of SARS-CoV-2 variants using genomic surveillance: insights from aircraft wastewater and nasal swabs at Kigali International Airport, Rwanda. IJID Reg. 2025;16:100678. doi:10.1016/j.ijregi.2025.100678

21. Suthar MS, Manning KE, Ellis ML, et al. The KP.2-adapted COVID-19 vaccine improves neutralising activity against the XEC variant. Lancet Infect Dis. 2025;25(3):e122–e123. doi:10.1016/S1473-3099(25)00007-6

22. Launching GLOWACON: A global initiative for wastewater surveillance for public health - European Commission. March 21, 2024. Accessed October 1, 2024. https://health.ec.europa.eu/latest-updates/launching-glowacon-global-initiative-wastewater-surveillance-public-health-2024-03-21_en

23. Liu P, Xu J. Genomic surveillance of SARS-CoV-2 in mainland China after ending the zero-COVID policy, December 2022-January 2023. J Infect. 2023;86(4):e84–e86. doi:10.1016/j.jinf.2023.02.040

24. Pan Y, Wang L, Feng Z, et al. Characterisation of SARS-CoV-2 variants in Beijing during 2022: an epidemiological and phylogenetic analysis. Lancet Lond Engl. 2023;401(10377):664–672. doi:10.1016/S0140-6736(23)00129-0

25. Wang S, Niu P, Su Q, et al. Genomic Surveillance for SARS-CoV-2 - China, September 26, 2022 to January 29, 2023. China CDC Wkly. 2023;5(7):143–151. doi:10.46234/ccdcw2023.026

26. Edens C, Clopper BR, DeVies J, et al. Notes from the Field: Reemergence of Mycoplasma pneumoniae Infections in Children and Adolescents After the COVID-19 Pandemic, United States, 2018-2024. MMWR Morb Mortal Wkly Rep. 2024;73(7):149–151. doi:10.15585/mmwr.mm7307a3

