## Supplementary Methods for "Traveler-based Genomic Surveillance: A Scalable Approach to Early Pathogen Detection and Global Biosecurity"

**Supplemental Methods**

***Program sites and recruitment***

TGS began at John F. Kennedy International Airport (JFK), Newark Liberty International Airport (EWR), and San Francisco International Airport (SFO) in September 2021 and later expanded to additional major U.S. international airports, including Hartsfield-Jackson Atlanta International Airport (ATL); Dulles International Airport (IAD) outside Washington, D.C; Seattle-Tacoma International Airport (SEA), Los Angeles International Airport (LAX), and Miami International Airport (MIA). Collection at ATL ended in July 2023.

Recruitment occurred post-immigration and customs.

Travelers provided consent, self-collected nasal swabs, and completed a questionnaire. Questionnaire updates included transition from paper to digital format, alignment with CDC information-collection standards, and the addition of a Spanish version. Travelers selected their travel origin from a list adapted from the International Organization for Standardization (ISO) standard 3166 (<https://www.iso.org/iso-3166-country-codes.html>). Forty-eight percent of staff who recruited volunteer travelers in the airport spoke a second language in addition to English. As a participation incentive, travelers received a COVID-19 antigen self-test kit.

***Sample handling and testing workflow***

Samples were shipped daily under cold chain to a central laboratory.

*Pooled testing phases*

- Sept 2021–Dec 2022**:** One swab per participant pooled by flight-origin country (5–25 swabs per pool).
- Dec 2022 onward**:** Two swabs collected; one pooled (5–10 swabs), one reserved for reflex testing.

Initially, participants were also given a mail-in SARS-CoV-2 saliva test kit for use 3–5 days later. In September 2022, follow-up saliva testing was discontinued when participation became anonymous.

Nasal swab samples were heat-inactivated at 70°C for 30 minutes, and total nucleic acids extracted using the MagMAX Viral/Pathogen II Nucleic Acid Isolation Kit (Thermo Fisher). Extracts were tested by qPCR using the TrueMark™ SARS-CoV-2/Flu A/Flu B/RSV Select Panel (Thermo Fisher). All nasal samples were biobanked for ≥30 days.

*Additional pathogen testing*

During December 5, 2023–March 16, 2024, samples from IAD and SFO were screened for *Mycoplasma pneumoniae* using the ThermoFisher TrueMark Respiratory Panel OpenArray. *M. pneumoniae*–positive samples were sent to CDC Division of Bacterial Diseases for confirmatory testing and macrolide susceptibility genotyping.^1,2^

***Wastewater sample collection and testing***

Airplane wastewater was collected from flights arriving from one Eastern Mediterranean country starting in February 2023 at JFK and December 2023 at IAD. Approximately 1 L of lavatory content was collected using a custom device that attached to the lavatory service panel port and the lavatory truck hose during routine maintenance.^3^ Triturator wastewater composite 24-hour samples were collected at SFO starting April 2023, Boston Logan International Airport (BOS) starting November 2023, and JFK starting August 2024.

Samples were shipped to a central laboratory for processing. Wastewater samples were concentrated with Nanotrap Microbiome Particles and extracted using a MagMAX Microbiome Ultra Nucleic Acid Isolation Kit (Thermo Fisher). Molecular testing for SARS-CoV-2 using RT-PCR for SARS-CoV-2 (February–December 2023) or RT-dPCR (December 2023–August 2024) with custom multiplexed panels for SARS-CoV-2, adenovirus F, RSV, influenza A and B, non-variola orthopoxvirus (NVO), norovirus GI, and norovirus GII.

***Sequencing and bioinformatics***

Sequencing was performed on SARS-CoV-2–positive nasal and wastewater samples and on influenza virus- and RSV-positive individual nasal samples. For targeted SARS-CoV-2 sequencing, purified total nucleic acid (TNA) was amplified by multiplex RT-PCR (NEB LunaScript Multiplex One-Step RT-PCR) using the ARTIC primer set (IDT ARTIC V5.3.2 NCOV-2019 Panel).^20^ Amplicons were pooled, and libraries prepared with a modified Illumina DNA Prep protocol. Libraries were sequenced on an Illumina NovaSeq 6000, targeting ~1.5 million paired-end 150 bp reads per sample. For influenza and RSV targeted enrichment sequencing, extracts were prepared using the Illumina RNA for Enrichment Sample Preparation Kit with the Respiratory Virus Oligo Panel, following the manufacturer’s protocol (Illumina, San Diego, CA). Libraries were validated for size and adapter dimer removal on an Agilent TapeStation 4200 D1000 High Sensitivity assay, quantified and normalized using the dsDNA High Sensitivity Assay on a Qubit 3.0 (Life Technologies, Carlsbad, CA), denatured/diluted, and pooled per manufacturer’s instructions. Pooled libraries were sequenced on the Illumina NovaSeq 6000 (v1.5 chemistry) to a target depth of ~20 million paired-end 150 bp reads per sample. Data was processed as previously described^4^ Genomes with >70% genome coverage at 10X read depth were retained. Lineages were assigned using Pangolin (SARS-CoV-2) or were otherwise subtyped.^5,6^ Wastewater lineage deconvolution was performed using Freyja.

Consensus genomes from nasal samples were submitted to [National Center for Biotechnology Information (NCBI) GenBank](https://www.ncbi.nlm.nih.gov/bioproject/989177) (all viruses) and Global Initiative on Sharing All Influenza Data (GISAID) (SARS-CoV-2). Raw SARS-CoV-2 sequencing reads for nasal and wastewater samples were submitted to NCBI Sequence Read Archive (SRA).

***Data analysis details***

WHO regional assignments used either flight-arrival origin or full itinerary. Taiwan was assigned to the Western Pacific region. SARS-CoV-2 sublineages (e.g., XBB*,* BA.2) were grouped for visualization. Wastewater and nasal swab results were analyzed separately but interpreted jointly to identify contemporaneous variant trends.

**Supplemental References**

1.	Wolff BJ, Benitez AJ, Desai HP, Morrison SS, Diaz MH, Winchell JM. Development of a multiplex taqMan real-time PCR assay for typing of Mycoplasma pneumoniae based on type-specific indels identified through whole genome sequencing. Diagn Microbiol Infect Dis. 2017 Mar;87(3):203-206. doi: 10.1016/j.diagmicrobio.2016.11.013. Epub 2016 Nov 25. PMID: 27923522.

2.	Wolff BJ, Thacker WL, Schwartz SB, Winchell JM 2008. Detection of Macrolide Resistance in *Mycoplasma pneumoniae* by Real-Time PCR and High-Resolution Melt Analysis. Antimicrob Agents Chemother 52:.<https://doi.org/10.1128/aac.00582-08>

3.	Morfino RC, Bart SM, Franklin A, et al. Notes from the Field: Aircraft Wastewater Surveillance for Early Detection of SARS-CoV-2 Variants - John F. Kennedy International Airport, New York City, August-September 2022. *MMWR Morb Mortal Wkly Rep*. 2023;72(8):210-211. doi:10.15585/mmwr.mm7208a3

4. Gratalo D, Friedman CR, Morley VJ, Qiu X, Rothstein AP, Tiburcio PB, et al. Environmental air monitoring in international airports: A novel approach for enhanced pathogen surveillance. medRxiv. 2025 Jan 1;2025.09.22.25336185. doi:10.1101/2025.09.22.25336185

5.	Rambaut A, Holmes EC, O’Toole Á, et al. A dynamic nomenclature proposal for SARS-CoV-2 lineages to assist genomic epidemiology. *Nat Microbiol*. 2020;5(11):1403-1407. doi:10.1038/s41564-020-0770-5

6.	Aksamentov I, Roemer C, Hodcroft EB, Neher RA. Nextclade: clade assignment, mutation calling and quality control for viral genomes. *J Open Source Softw*. 2021;6(67):3773. doi:10.21105/joss.03773
