## Supplementary material for "Traveler-based Genomic Surveillance: A Scalable Approach to Early Pathogen Detection and Global Biosecurity": Figures

**Figure 1**. Nasal sample collection and testing strategy for arriving international air travelers, program airports, and weekly participation over time — Traveler-based Genomic Surveillance program, United States, September 2021–August 2024

Abbreviations: RSV: respiratory syncytial virus, JFK: John F. Kennedy International Airport, EWR: Newark Liberty International Airport, SFO: San Francisco International Airport, ATL: Hartsfield-Jackson Atlanta International Airport, IAD: Dulles International Airport, SEA: Seattle-Tacoma International Airport, LAX: Los Angeles International Airport, MIA: Miami International Airport

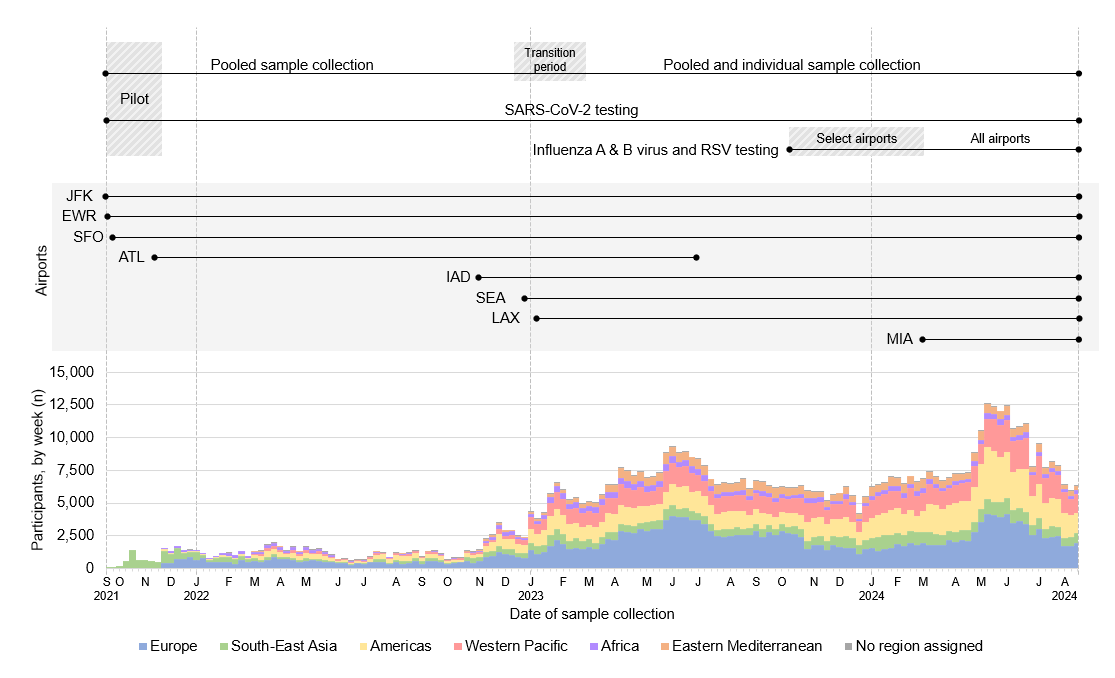

**Supplementary Figure 1**. Nasal swab sampling participation of arriving international air travelers, by country of travel origin — Traveler-based Genomic Surveillance program, United States, September 2021–August 2024

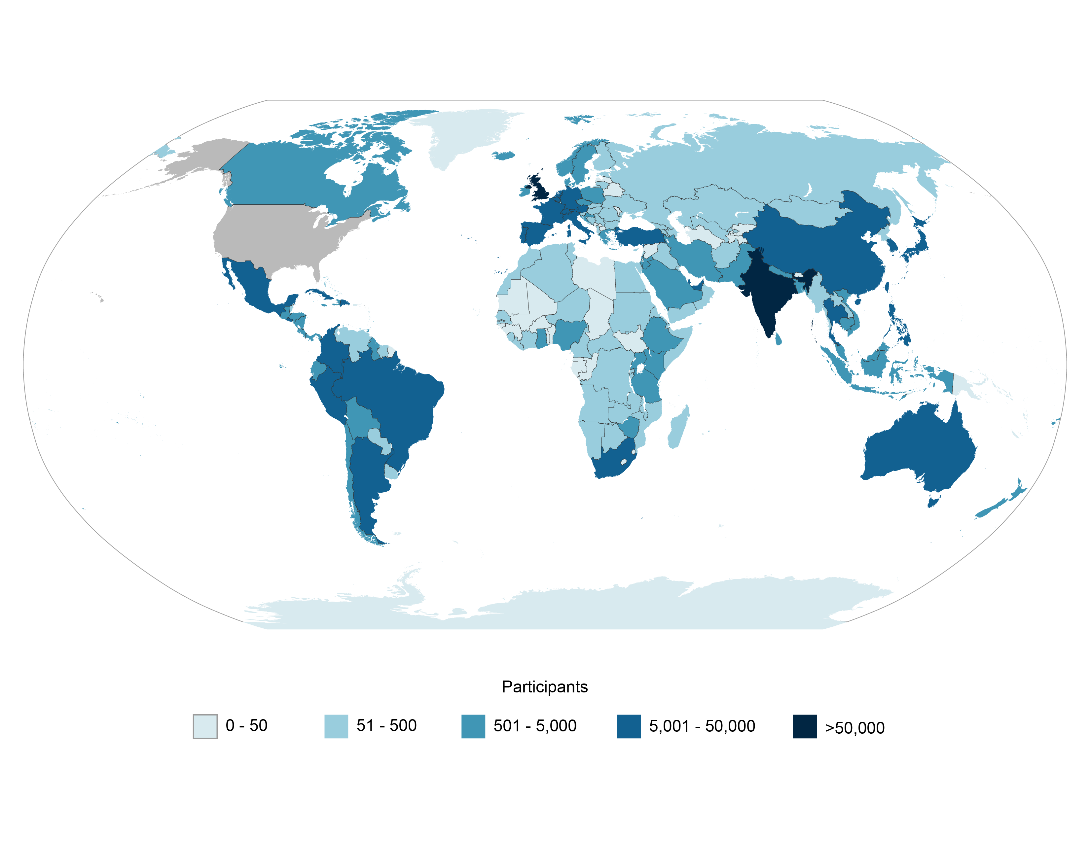

**Figure 2**. SARS-CoV-2 test and sequencing results for arriving international air traveler nasal samples, by week — Traveler-based Genomic Surveillance program, United States, September 2021–August 2024. (**A**) Percentage of positive SARS-CoV-2 test results for pooled nasal samples (black) and individual nasal samples (blue inset) from participating travelers. (**B**) Percentage of SARS-CoV-2 genomic sequences assigned to major Pangolin lineage groups from pooled nasal samples from participating travelers. Because of sample size limitations and methodologic changes during the pilot, graphics show results beginning November 28, 2021.

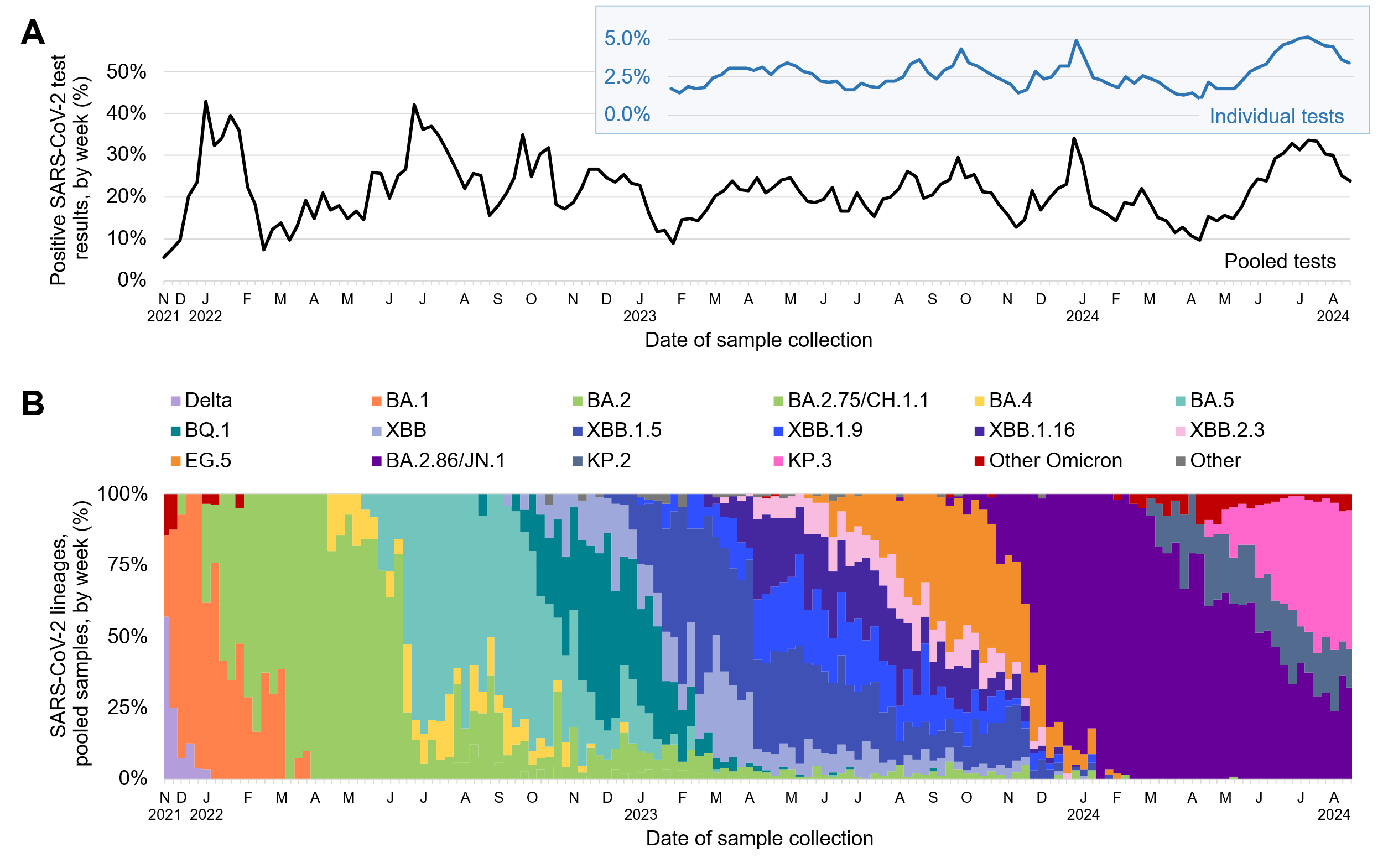

**Supplementary Figure 2**. Test results and participation for pooled nasal tests and follow-up saliva tests of arriving international air travelers — Traveler-based Genomic Surveillance program, United States, September 2021–September 2022. (**A**) Percentage of positive SARS-CoV-2 test results for pooled nasal samples (black) and follow-up saliva tests (orange) from arriving travelers, excluding results from the pilot period because of subsequent methodological changes. (**B**) Number of travelers participating in follow-up saliva testing after nasal testing participation.

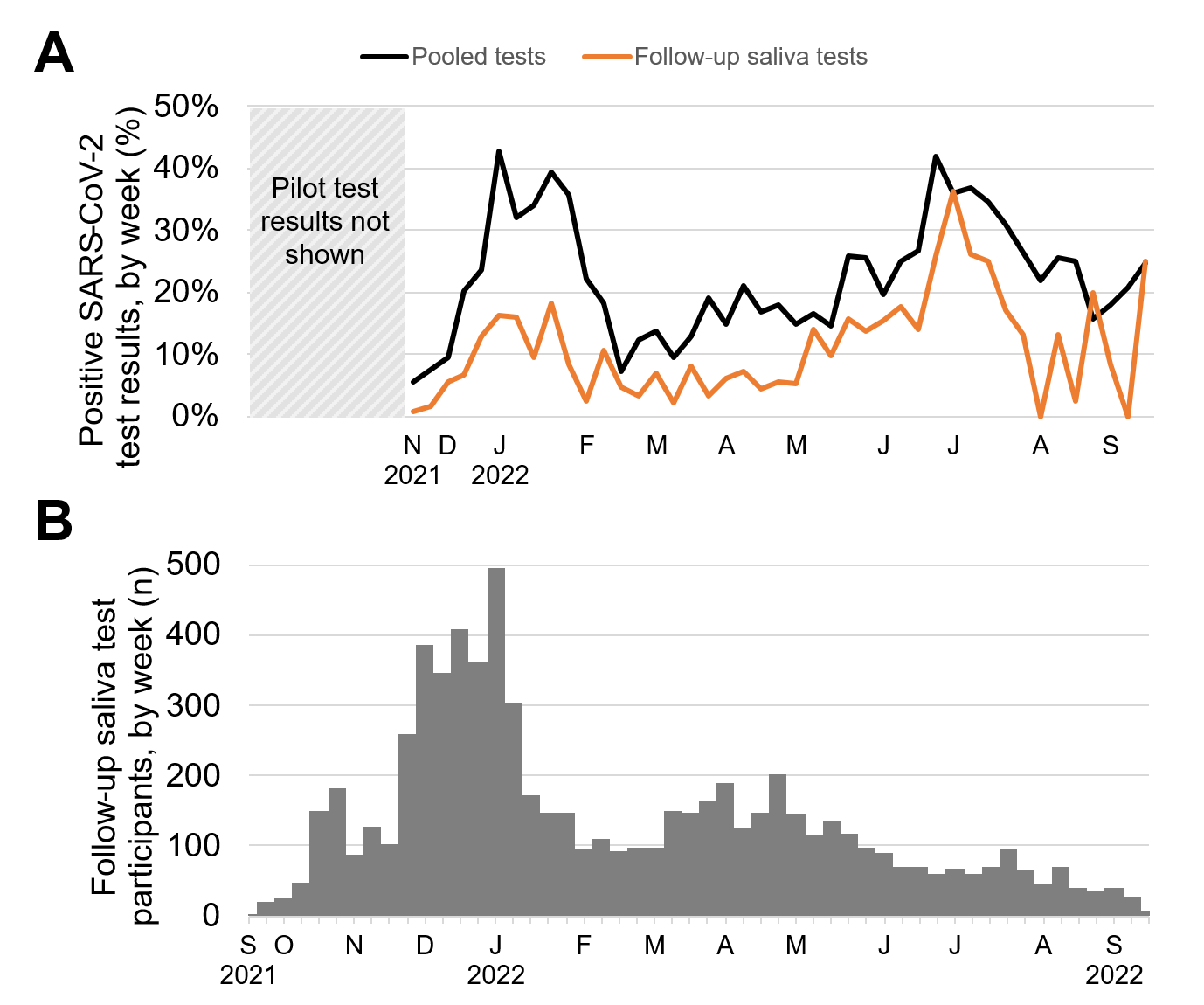

**Supplementary Figure 3**. SARS-CoV-2 test and sequencing results for arriving international air traveler nasal samples, by week and World Health organization (WHO) region — Traveler-based Genomic Surveillance program, United States, January 2023–August 2024. (**A–F**) Percentage of positive SARS-CoV-2 test results for individual nasal samples from participating travelers originating in each WHO region. (**G–L**) Number of SARS-CoV-2 genomic sequences assigned to major Pangolin lineage groups in individual nasal samples from participating travelers.

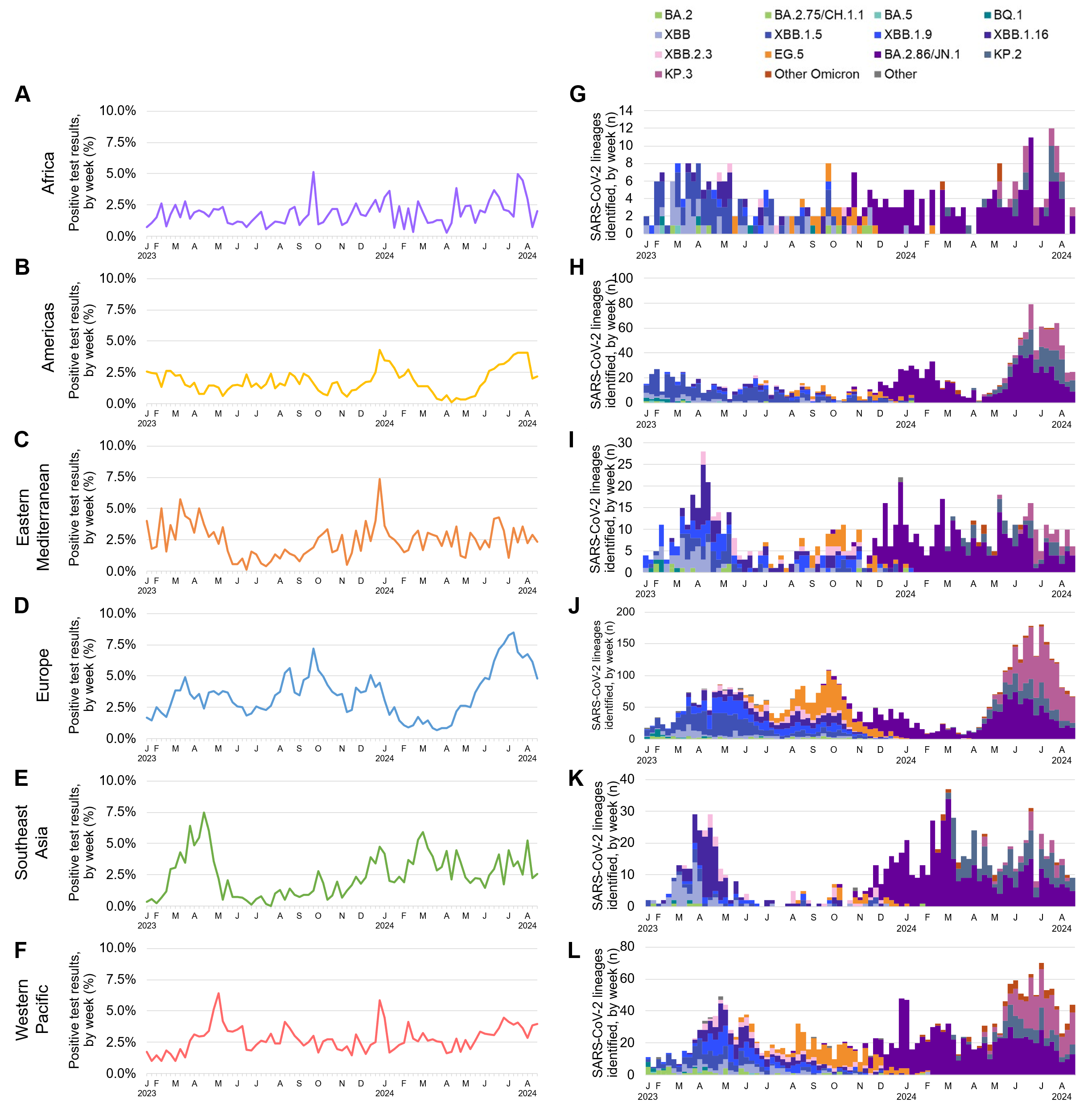

**Figure 3**. Influenza A virus, influenza B virus, and RSV test and sequencing results in nasal samples of arriving international air travelers, by week — Traveler-based Genomic Surveillance program, United States, October 2023–August 2024. (**A**) Percentage of positive influenza A virus, influenza B virus, and RSV test results for individual nasal samples from participating travelers. (**B–D**) Influenza A virus subtypes (**B**), influenza B virus subtypes (**C**), and RSV genotypes (**D**) for viral genomic sequences from individual nasal samples from participating travelers.

Abbreviations: RSV: respiratory syncytial virus

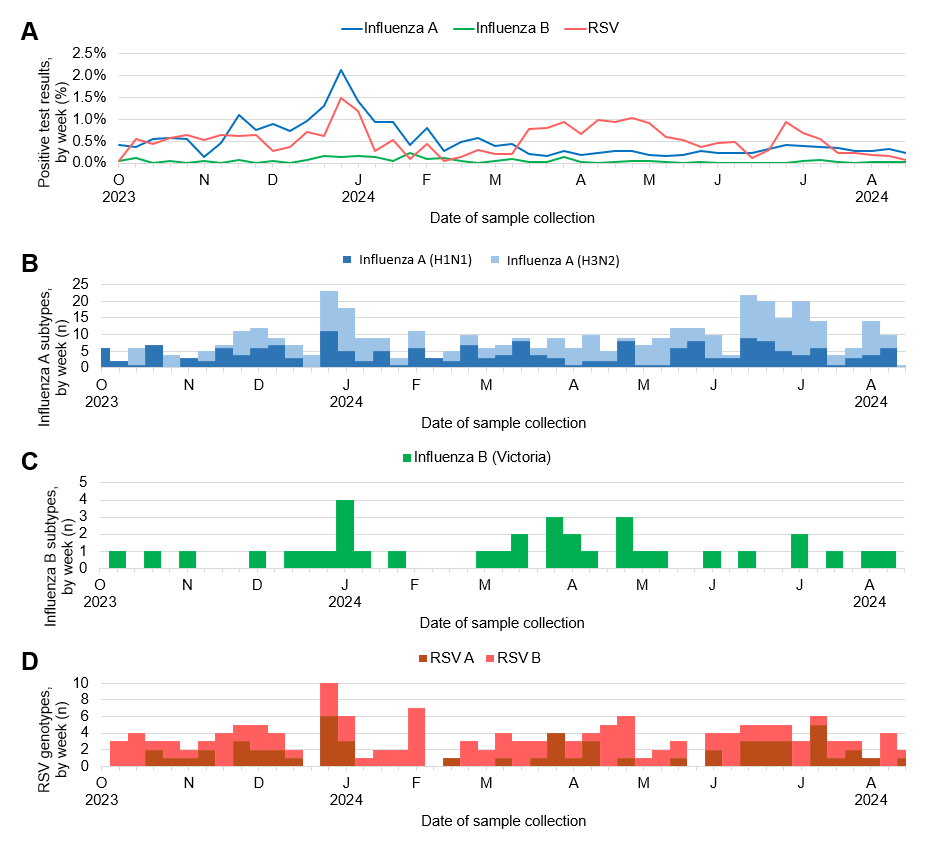

**Figure 4**. Sample collection and SARS-CoV-2 test results for airplane and triturator wastewater samples, by week — Traveler-based Genomic Surveillance program, United States, February 2023–August 2024. The triturator is a drain where waste from multiple flights is discharged and homogenized before later being combined with terminal wastewater. (**A**) Number of wastewater samples collected, by airport and wastewater collection type. SFO and BOS triturator sampling was suspended in June and July 2024 for operational issues. (**B**) Percentage of positive SARS-CoV-2 test results for airplane (blue) and triturator (orange) wastewater samples. (**C–D**) Percentage of SARS-CoV-2 genomic sequences assigned to major Pangolin lineage groups from airplane (**C**) and triturator (**D**) wastewater samples. Airplane wastewater samples were collected from flights arriving from the Eastern Mediterranean region. Gaps indicate weeks when no samples were tested or when no lineages were identified.

Abbreviations: JFK: John F. Kennedy International Airport, IAD: Dulles International Airport, SFO: San Francisco International Airport, BOS: Boston Logan International Airport

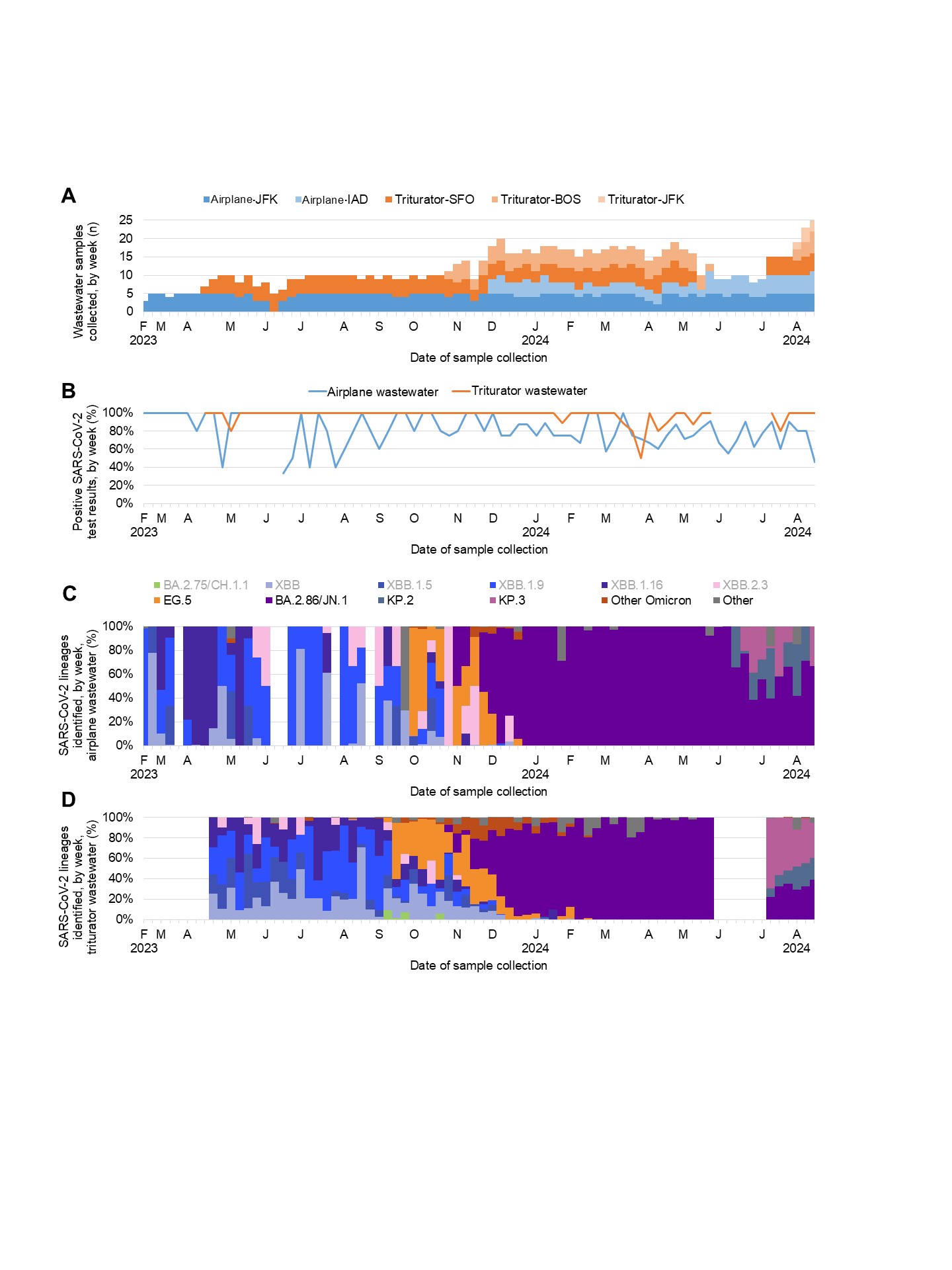

**Figure 5**. Adenovirus F, respiratory syncytial virus, influenza A virus, influenza B virus, norovirus GI, norovirus GII, and non-variola orthopoxvirus test results for airplane (**A–G**) and triturator wastewater (**H–N**) samples, by week — Traveler-based Genomic Surveillance program, United States, December 2023–August 2024. Airplane wastewater samples were collected from flights arriving from the Eastern Mediterranean region. The triturator is a drain where waste from multiple flights is discharged and homogenized before later being combined with terminal wastewater. The non-variola orthopoxvirus test detects certain poxviruses including Monkeypox virus. Gaps indicate weeks when no samples were tested.

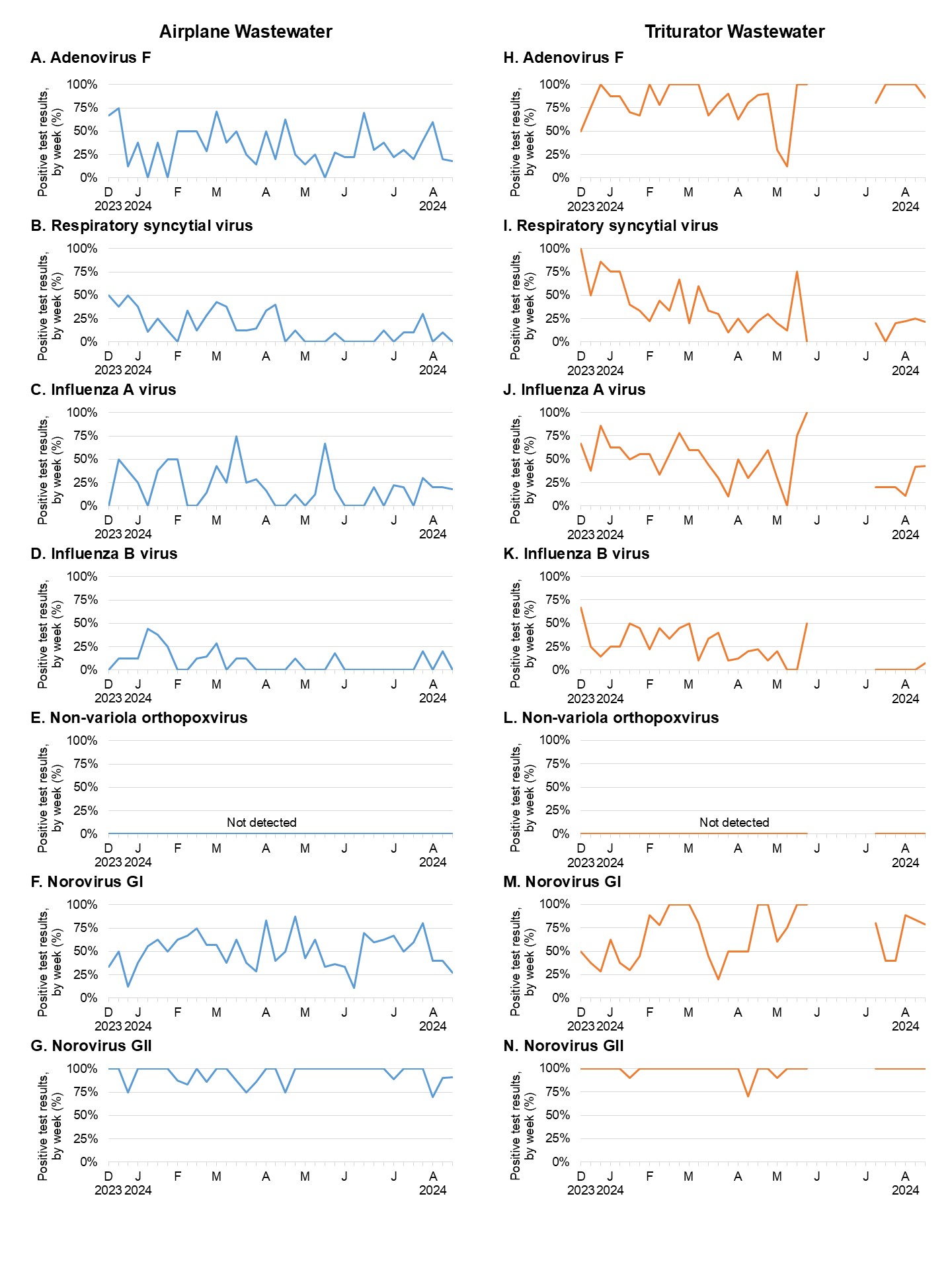

**Figure 6**. Flight route coverage, weekly nasal sampling results from arriving air travelers originating from China, weekly China COVID-19 epidemiology, and weekly program participation by travelers from China during 2022–2023 China COVID-19 surge — Traveler-based Genomic Surveillance (TGS) program, United States, November 2022–February 2023. (**A**) Map showing flight routes from cities in China (orange) and other countries in East Asia (blue) to TGS program airports. (**B**) Number of TGS participants originating in mainland China (excluding Hong Kong and Macau Special Administrative Regions). TGS program actions to increase participation are labeled. (**C**) Comparison of number of COVID-19 cases reported by China to the World Health Organization (red, https://data.who.int/dashboards/covid19/cases) with percentage of positive SARS-CoV-2 tests in nasal samples from participating travelers originating in China (blue). A pre-boarding requirement for proof of a negative test or recent recovery from COVID-19 was in place for U.S-bound air passengers from China during January 5, 2023–March 10, 2023.

Abbreviations:

SEA: Seattle-Tacoma International Airport,, LAX: Los Angeles International Airport, SFO: San Francisco International Airport, EWR: Newark Liberty International Airport, JFK: John F. Kennedy International Airport, IAD: Washington Dulles International Airport, ATL: Hartsfield-Jackson Atlanta International Airport

PEK: Beijing Capital International Airport, PVG: Shanghai Pudong International Airport, XMN: Xiamen Gaoqi International Airport, CAN: Guangzhou Baiyun International Airport, SZX: Shenzhen Bao'an International Airport, HKG: Hong Kong International Airport, TPE: Taiwan Taoyuan International Airport, ICN: Incheon International Airport, NRT: Narita International Airport, HND: Tokyo International Airport, KIX: Kansai International Airport, SGN: Tan Son Nhat International Airport, SIN: Singapore Changi Airport.

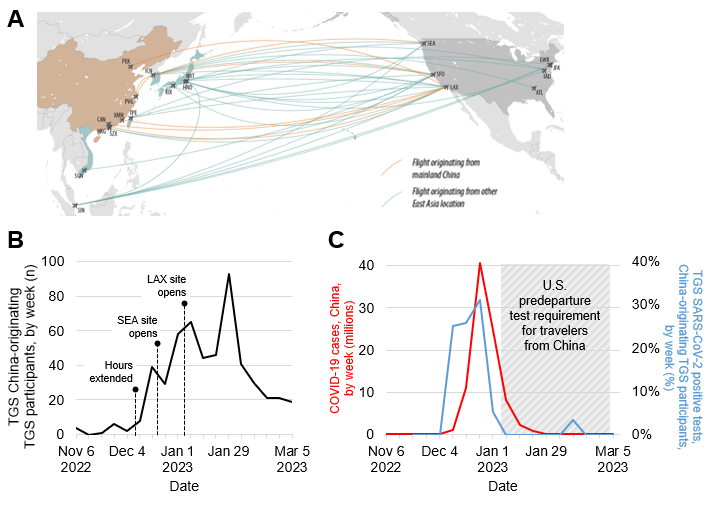
