## Supplementary material for "Traveler-based Genomic Surveillance: A Scalable Approach to Early Pathogen Detection and Global Biosecurity": Tables

**Supplemental Table 1**. Participant responses to demographic and travel history questions, by date of participation — Traveler-based Genomic Surveillance Program, United States, September 2021–August 2024.

|  | **Date Range^a^** | **Questions and Responses** | **n** | **% of respondents** |
| --- | --- | --- | --- | --- |
| September 29, 2021– August 25, 2024 | | **All participants** | **694,798** |  |
| **Traveler origin region** | | | | |
| September 29, 2021–January 21, 2023 | | **Origin based on origin of flight leg arriving in the United States** | **88,481** |  |
|  |  | Africa | 7,273 | 8.2% |
|  |  | Americas | 16,414 | 18.6% |
|  |  | Eastern Mediterranean | 1,149 | 1.3% |
|  |  | Europe | 36,067 | 40.8% |
|  |  | South-East Asia | 20,498 | 23.2% |
|  |  | Western Pacific | 6,709 | 7.6% |
|  |  | Unknown | 371 | 0.4% |
| December 11, 2022–  August 25, 2024 | | **Origin based on origin country of the traveler’s self-reported flight itinerary** | **606,317** |  |
|  |  | Africa | 30,448 | 5.0% |
|  |  | Americas | 143,419 | 23.7% |
|  |  | Eastern Mediterranean | 45,699 | 7.5% |
|  |  | Europe | 203,962 | 33.6% |
|  |  | South-East Asia | 61,663 | 10.2% |
|  |  | Western Pacific | 120,305 | 19.8% |
|  |  | Unknown | 821 | 0.1% |
| **Age** | | | | |
| September 29, 2021– August 25, 2024 | | **Participants who were asked their date of birth or age range** | **694,798** |  |
|  |  | **Respondents** | **630,621** |  |
|  |  | 18–49 years | 419,938 | 66.6% |
|  |  | 50–64 years | 158,232 | 25.1% |
|  |  | 65+ years | 39,698 | 6.3% |
|  |  | Prefer not to answer | 12,753 | 2.0% |
| **Country of residence** | | | | |
| September 29, 2021-August 25, 2024 | | **Participants who were asked their country of residence** | **694,798** |  |
|  |  | **Respondents** | **603,651^b^** |  |
|  |  | United States | 352,360 | **58.4%** |
|  |  | Outside the United States | 243,729 | **40.4%** |
|  |  | Prefer not to answer | 7,562 | **1.2%** |
| **Sex** | | | | |
| September 29, 2021-August 25, 2024 | | **Participants who were asked their sex** | **694,798** |  |
|  |  | **Respondents** | **647,746^b^** |  |
|  |  | Female | 327,325 | 50.5% |
|  |  | Male | 306,774 | 47.4% |
|  |  | Unavailable^c^ | 13,647 | 2.1% |
|  | **Race/Ethnicity** | | | |
|  | September 29, 2021–June 1, 2024 | **Participants who were asked one multiple-response race question and one ethnicity question** | **590,244** |  |
|  |  | **Respondents: Race^d^** | **513,249^b^** |  |
|  |  | American Indian or Alaska Native | 13,279 | 2.6% |
|  |  | Asian | 138,785 | 27.0% |
|  |  | Black or African American | 52,930 | 10.3% |
|  |  | Native Hawaiian or Other Pacific Islander | 4,184 | 0.8% |
|  |  | White | 266,907 | 52.0% |
|  |  | Prefer not to answer | 47,121 | 9.2% |
|  |  | **Respondents: Ethnicity** | **462,385^b^** |  |
|  |  | Hispanic or Latino | 85,151 | 18.4% |
|  |  | Not Hispanic or Latino | 351,733 | 76.1% |
|  |  | Prefer not to answer | 25,501 | 5.5% |
|  | June 2, 2024–  August 25, 2024 | **Participants who were asked one multiple-response race/ethnicity question^d^** | **104,554** |  |
|  |  | **Respondents** | **103,115** |  |
|  |  | American Indian or Alaska Native | 1,958 | 1.9% |
|  |  | Asian | 26,789 | 26.0% |
|  |  | Black or African American | 6,657 | 6.5% |
|  |  | Hispanic or Latino | 19,081 | 18.5% |
|  |  | Middle Eastern or North African | 1,957 | 1.9% |
|  |  | Native Hawaiian or Other Pacific Islander | 899 | 0.9% |
|  |  | White | 46,354 | 45.0% |
|  |  | Prefer not to answer | 2,650 | 2.6% |

^a^ Different versions of some questions were employed throughout the analysis period. Date ranges sometimes overlap because of phased implementation of new questionnaires by airport.

^b^ In some early versions of the questionnaire, no affirmative “Prefer not to answer” option was given.

^c^ Includes affirmative “Prefer not to answer” responses and any responses other than male or female.

^d^ Summed percentages may exceed 100%; participants could choose multiple options.
